# Increasing Trust in Real-World Evidence Through Evaluation of Observational Data Quality

**DOI:** 10.1101/2021.03.25.21254341

**Authors:** Clair Blacketer, Frank J Defalco, Patrick B Ryan, Peter R Rijnbeek

**Affiliations:** Observational Health Data Analytics, Janssen Research and Development, LLC, Titusville, NJ, USA; Department of Medical Informatics, Erasmus University Medical Center, Rotterdam, The Netherlands; Department of Biomedical Informatics, Columbia University, New York, USA

## Abstract

Advances in standardization of observational healthcare data have enabled methodological breakthroughs, rapid global collaboration, and generation of real-world evidence to improve patient outcomes. Standardizations in data structure, such as use of Common Data Models (CDM), need to be coupled with standardized approaches for data quality assessment. To ensure confidence in real-world evidence generated from the analysis of real-world data, one must first have confidence in the data itself. The Data Quality Dashboard is an open-source R package that reports potential quality issues in an OMOP CDM instance through the systematic execution and summarization of over 3,300 configurable data quality checks. We describe the implementation of check types across a data quality framework of conformance, completeness, plausibility, with both verification and validation. We illustrate how data quality checks, paired with decision thresholds, can be configured to customize data quality reporting across a range of observational health data sources. We discuss how data quality reporting can become part of the overall real-world evidence generation and dissemination process to promote transparency and build confidence in the resulting output. Transparently communicating how well CDM standardized databases adhere to a set of quality measures adds a crucial piece that is currently missing from observational research. Assessing and improving the quality of our data will inherently improve the quality of the evidence we generate.

## INTRODUCTION

As the amount of observational health data available to researchers continues to grow, regulatory agencies like the U.S. Food and Drug Administration (FDA)[1] and the European Medicines Agency (EMA)[2] have seen the value of using real-world evidence but there is still some concern and lack of trust in real-world data. Use of standardized data structures like the OMOP Common Data Model (CDM)[3] have allayed some of these fears but standardization alone is not enough. Rigorous data quality assessments are needed to evaluate the quality of data with which evidence is generated.

The use of the OMOP CDM has lain the groundwork for impressive methodological and clinical breakthroughs in the areas of population-level effect estimation, patient-level prediction, and clinical characterization.[4–6] The recent *Large-Scale Evidence Generation and Evaluation across a Network of Databases* (LEGEND) study published in the Lancet is an exemplar of these ideas as the authors not only delivered relevant information about first-line antihypertensive drugs but also a novel approach to generating evidence using a systematic framework.[7] However, in order for regulatory agencies and clinicians to make decisions using such evidence, there needs to be trust not only in the methodologies employed but the underlying data itself.

The pitfalls of the secondary use of observational data to support research are well documented.[8–13] Typically these data are collected either for billing or diagnostic purposes and not with research endpoints in mind. Von Lucadou, et al. notes that information in the electronic health record may not be as granular as data captured during the course of a clinical trial and time stamps of clinical events should be examined prior to inferring temporal relationships.[9] Additionally, it is often the case that clinical ideas are captured in free-text fields, as described by Varela, et al.[11] It can be easy to overlook the multiple ways a white blood cell count is recorded, for example. One physician might use “WBC” while another uses “White Blood Cell” and yet another “White BC”. These types of inconsistencies can lead to misclassification and measurement error.

Such issues are concerning but not unknown to clinical research networks (CRNs). The Sentinel Initiative,[14,15] the National Patient-Centered Clinical Research Network (PCORnet®),[16,17] and the Pediatric Learning Health System (PEDSnet),[18,19] among others, have all built processes and tools meant to identify data quality problems well in advance of any analytics that might be performed using the data. Historically, OHDSI promoted a tool known as the Automated Characterization of Health Information at Large-scale Longitudinal Evidence System (ACHILLES) to assess the quality of data in the OMOP CDM format.[20] ACHILLES is primarily used for database characterization. It computes a set of aggregate summary statistics such as gender and age stratifications of persons included, average follow-up time, and distribution of diagnosis codes, among others. These statistics are then assessed for quality by running the ACHILLES Heel rules against the aggregated summaries rather than on the database itself. These rules include looking for patients with an age less than zero and prescription dispensing records with implausible drug quantities.

A comparison of the data quality assessment (DQA) programs across six different CRNs in 2017 revealed that OHDSI had the fewest data quality checks in place (172) while the other networks ranged from 875 up to 3,434 checks.[21] The reason for this difference is that OHDSI as an open collaborative has traditionally left DQA to the individual data owners. Lead protocol investigators are responsible for making a “fitness-for-use” decision and independently determine if a dataset is suitable to answer a clinical question. As OHDSI continues to move in the direction of large-scale network research[22–24] a more robust data quality tool is needed to ensure that a participant’s data comply with community defined standards.

It is our goal to address the need for better data quality processes with the development of the Data Quality Dashboard (DQD). In this paper we describe the methods used to design the tool through community engagement. Next, we describe the inner workings of the DQD and run it against a US claims database as a proof-of-concept to show utility in practice. Finally, we discuss future enhancements and the potential for this tool to change the way observational data is utilized within OHDSI and beyond.

## BACKGROUND

To take advantage of the tools and methodologies[4,25] available to the OHDSI community, collaborators must first convert their data to the OMOP CDM. Data owners, clinicians, and OMOP experts all come together to standardize a source database by putting it in the structure of the model and applying agreed upon conventions through a process known as extract, transform, load (ETL). This is the expected function of any standard data model but where OMOP differs is that not only the structure but also the content of the data is standardized, harmonizing on the SNOMED vocabulary[26] for conditions and RxNorm vocabulary[27] for drugs, for example. It is this semantic standardization that facilitates international adoption and fosters rapid collaboration.

Once a database is converted to the CDM ideally it should be assessed for quality prior to using it for generating evidence. Some collaborators, like PEDSNet, designed their own data quality tools to keep track of metrics they were most interested in.[19] For many collaborators, the ACHILLES Heel report served this function up until now. The tool ran a set of checks against a CDM instance and reported them back to the user as an available option in the Achilles characterization package^1^. The Heel report gave an overview of potential data quality issues, but it did not allow the user to change how a pass or fail for a given check was determined, did not share the SQL query that was run to produce the result, nor did it give the option to capture any metadata about why a perceived quality issue might be occurring. There is also no process for extending the tool either by adding features or new data quality checks.

We therefore developed the DQD as a stand-alone R package to improve upon prior work and to fill in the gaps left by the Achilles Heel report. By focusing our attention to assessing data after the conversion to the OMOP CDM, the standardized structure and content directly enabled the creation of a standardized quality control framework with the DQD at the center (figure 1).

**Figure 1:**
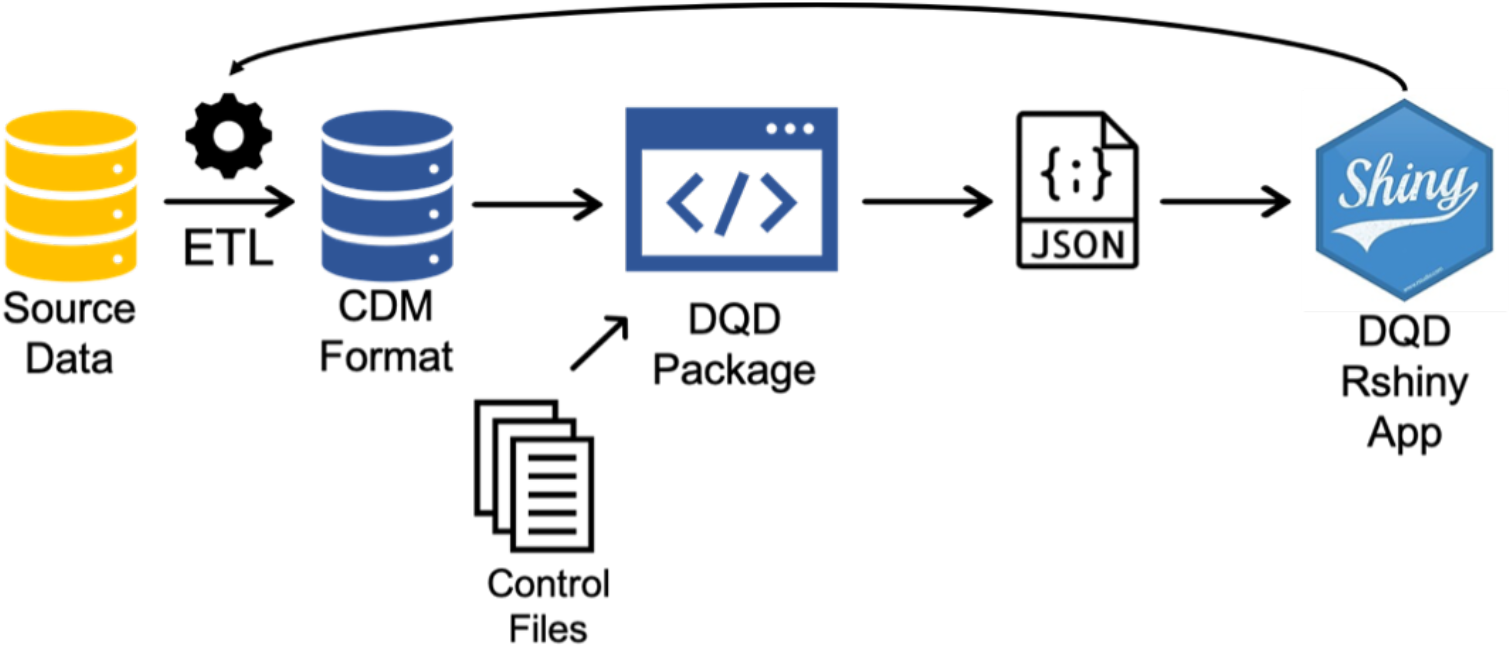
The standard quality control framework

## MATERIALS AND METHODS

To lead this initiative a few interested OHDSI collaborators formed a committee to identify high-level data quality issues important to the community. We aligned with the framework described by Kahn, et. al.[28] as a way to organize our approach. Kahn and his colleagues identified the categories of conformance, completeness, and plausibility into which most, if not all, data quality checks can be grouped.

Conformance checks measure how well a database conforms to specified formats and relational constraints. The committee applied this idea to the OMOP CDM, describing issues such as whether a CDM instance contains all required fields, if fields that are defined as primary keys contain unique values, and if a foreign key value is present in its corresponding primary key field.

Completeness checks look at the frequency of values in a given dataset without examination of the values themselves. In terms of the CDM the idea of completeness can likewise be used to understand the quality of vocabulary mapping. The committee identified this as an area of importance for an OMOP data quality solution, including checks to evaluate the proportion of source values (ICD10CM, CPT4, Read, etc.) that were not mapped to standard concepts.

Plausibility checks are meant to gauge the believability of values in a dataset. This can take many forms like making sure a person’s health care encounters all occur on or after their birthdate or looking to see that no one has a weight of zero kilograms recorded.

After deciding on the data quality checks to implement in a new data quality tool the committee turned their sights to the features they would like included. These were chosen based on discussions with key stakeholders from both academia and industry about their needs in this space. Considering the myriad of infrastructure constraints present in the community it should be a stand-alone application that any user can run out-of-the box. It should be easily scalable to allow inclusion of additional checks over time. It should also be flexible to allow adjustment of the checks and failure thresholds based on apriori knowledge of a database. The results of the data quality assessment should be easily shareable, in some form or fashion that is not a burden to the data owner.

With these requirements in mind, we devised a framework that would allow us to use the structure of the data model to our advantage. Since we already know the schema of every database that will run this tool, we didn’t have to focus on how to write and execute individual data quality checks; instead, we were able to abstract a layer and define data quality *ideas*. For example, what if it is important to assess the number of persons in the PERSON table that don’t have a record in the VISIT_OCCURRENCE table?

That check can be assessed using a simple SQL statement against an OMOP CDM instance:

> **SELECT** COUNT (DISTINCT P.person_id)
>
> **FROM** PERSON as P
>
> **LEFT JOIN** VISIT_OCCURRENCE as V
>
> **ON** P.person_id = V.person_id
>
> **WHERE** V.person_id IS NULL

As a data quality idea we are simply evaluating the degree to which persons in the PERSON table are represented in a fact table (VISIT_OCCURRENCE in this example). In the OMOP CDM the field PERSON.person_id is a primary key with corresponding foreign keys in all clinical fact tables. Using that constraint as our guide, the abstraction of this data quality check to a data quality idea results in a SQL statement like this:

> **SELECT** COUNT (DISTINCT P.person_id)
>
> **FROM** PERSON as P
>
> **LEFT JOIN** @cdmTable
>
> **ON** P.person_id = @cdmTable.person_id
>
> **WHERE** @cdmTable.person_id IS NULL

Where @cdmTable represents the universe of clinical fact tables that have a foreign key to the PERSON table. Each of them can be rotated into the SQL statement to take the place of the parameter. If there are 15 of such tables then the above SQL would automatically generate 15 data quality checks from one data quality idea.

Using the process described above as our foundation we developed the Data Quality Dashboard (DQD) R package that systematically runs and evaluates data quality checks based on the structure of the Common Data Model and pre-specified failure thresholds.

To test it, the DQD was run on the IBM Marketscan® Multi-State Medicaid (MDCD) database. This database contains adjudicated US health insurance claims for Medicaid enrollees from multiple states and includes hospital discharge diagnoses, outpatient diagnoses and procedures, and outpatient pharmacy claims as well as ethnicity and Medicare eligibility. The major data elements contained within this database are outpatient pharmacy dispensing claims, inpatient, and outpatient medical claims. The data does not contain laboratory results.

## RESULTS

In total, 20 high-level data quality ideas were identified: 5 completeness, 8 conformance, and 7 plausibility, the complete list of which is included in appendix 1. The example given above to assess the extent to which persons in the PERSON table are represented in the clinical fact tables became the data quality idea, or check type, *measurePersonCompleteness*. In terms of OMOP, completeness means not only missingness but vocabulary mapping completeness. The results of two check types, *standardConceptRecordCompleteness* (SCRC) and *sourceValueCompleteness* (SVC), work together to show how well the diagnostic, procedural, and drug codes, etc. (source values) in a database have been mapped to the standard terminology as defined by OMOP. SCRC counts the number of records for a given table that have been mapped to a CONCEPT_ID of zero where a Standard Concept is expected. If applied to the CONDITION_OCCURRENCE table, for example, it would count the number of records with a CONDITION_CONCEPT_ID of zero. In a situation where a source value in a database cannot be mapped to a Standard Concept, a zero is used to denote “No matching concept”. Therefore, if a large number of records in a table have a Standard Concept of zero, those records cannot be used in an analysis as standardized analytics rely on standardized vocabularies.

The SVC check type, on the other hand, quantifies the number of distinct source values in a database that have been mapped to zero. Using the example from earlier, the values “WBC”, “White Blood Cell”, and “White BC” are all ways in which the clinical idea of a white blood cell count measurement might be represented. These values are all free text and as such do not have an automatic mapping to a Standard Concept. Thus, records with these values would be given a Standard Concept Id of zero during the ETL conversion process. If database A has 5,000 records in the resulting MEASUREMENT table with these source values, then the SCRC check type applied to the MEASUREMENT_CONCEPT_ID field in the MEASUREMENT table would return 5,000 records mapped to zero while the SVC check type applied to the same field would return 3 distinct values mapped to zero. This can be interpreted to mean there are 5,000 records representing 3 distinct values that are missing a mapping to a Standard Concept in the MEASUREMENT_CONCEPT_ID field of database A. The relationship between these two checks will signal different vocabulary or ETL changes to be made. A high SCRC and high SVC might indicate that the database has a proprietary coding system not represented in the OMOP vocabulary. In such case either the coding system is added to the vocabulary or the source codes are manually mapped to standard concepts. A high SCRC and low SVC is often due to a small number of catch-all values like “unknown” or “UNK”. These do not have any real meaningful analytic use so the records are either ignored (by increasing the failure threshold) or they are removed. A low SCRC and high SVC usually means that the source values representing the highest number of records were mapped to standard concepts and the rest were mapped to zero. This is an accepted ETL practice as the records mapped to zero are retained for later use if necessary. A low SCRC and low SVC means that a large number of source codes representing a large number of records were mapped to standard concepts. This is the ideal scenario for an ETL though it is important to monitor these values over time in the event the source coding practices change.

The *plausibleTemporalAfter* check type is used to evaluate the temporal relationships between date values. Applied to the VISIT_OCCURRENCE table this check quantifies the number of visits with a visit end date prior to the visit start date. If such visits are found, the ETL must make a choice how to handle them. The most common practice is to choose either the start or end date as listed in the source data and use that value for both fields (VISIT_START_DATE, VISIT_END_DATE) in the standardized dataset. This allows for the information from the visit to be retained while eliminating the temporal inconsistency.

When the *plausibleTemporalAfter* check type is applied to the CONDITION_OCCURRENCE table it becomes a way to measure the number of condition records that occur prior to a person’s birth. There can be many reasons for records to be written with incorrect dates but usually these are patient history records where no date was given by the patient and, instead, a default date is assigned by the electronic health record. As it impossible to discern the correct date for these records the typical recourse is to remove them.

These check types were all written as parameterized SQL statements. To resolve and then run these SQL statements the DQD reads a set of included control files that detail each table and field in the CDM, their constraints, and their relationships to one another. These files also indicate which data quality check types should be run on which fields and what thresholds should be applied to the results to determine a pass or fail, all of which can be edited by the user. The tool then takes these files and swaps out the parameters in the SQL statements for the values indicated in the control files using the SqlRender R package.[29]

With this approach the 20 checks types are resolved to over 3,300 individual quality checks: 396 completeness, 779 conformance, and 2126 plausibility. The DQD then compiles all results into a JSON file as the default output, though the user can also specify the option to write the results back to a table in the database. The resulting JSON (or table) contains all information produced by the tool and is read by the package to render an interactive Rshiny[30] user interface (figure 1).

The first screen that greets the user is the overview tab, shown in figure 2 displaying the result of running the DQD on the MDCD database. The tables NOTE, NOTE_NLP, and SPECIMEN are not populated for MDCD, resulting in a lower total number of checks at 3,124 instead of 3,300.

**Figure 2:**
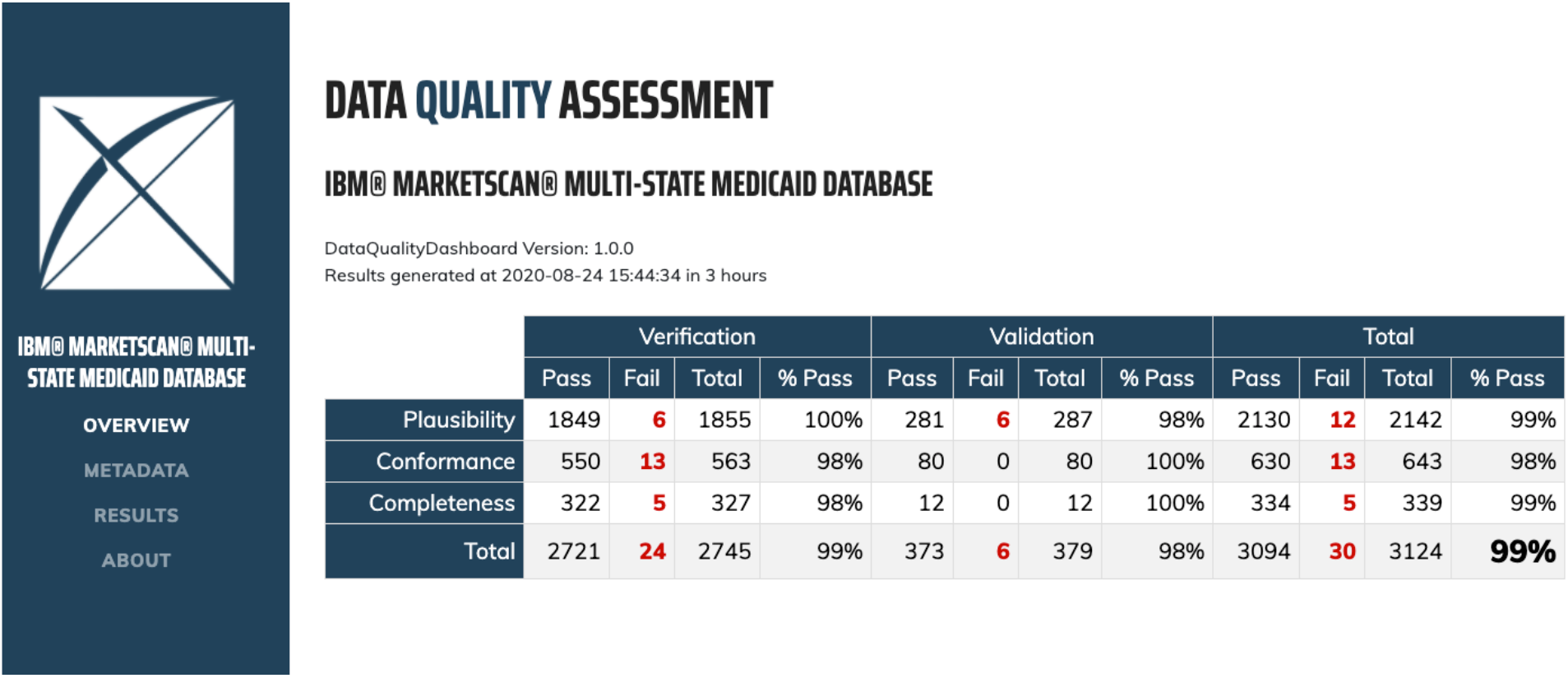
The overview tab of the DQD showing data quality check pass percentages by category in MDCD.

The overview tab gives a high-level summary of the total number of passes and failures by Kahn category. In figure 2, the database examined has 13 conformance failures, or instances where it does not conform to the specifications of the OMOP CDM. It has 5 completeness failures related to potentially missing data and 12 plausibility failures which could be a myriad of issues including incorrect dates or implausible measurement values. To explore each of these failures the results tab (figure 3) shows one line per check run. Across the top is the option to filter by pass/fail status, CDM table, and Kahn context. The plus sign on each line expands to show the exact SQL query that was run to achieve that result, allowing the user to pinpoint the identified failing records in their dataset. A publicly available instance of these data quality results can be found at https://data.ohdsi.org/DataQualityDashboardMDCD/ and the JSON file with the full result set is available in supplementary materials.

**Figure 3:**
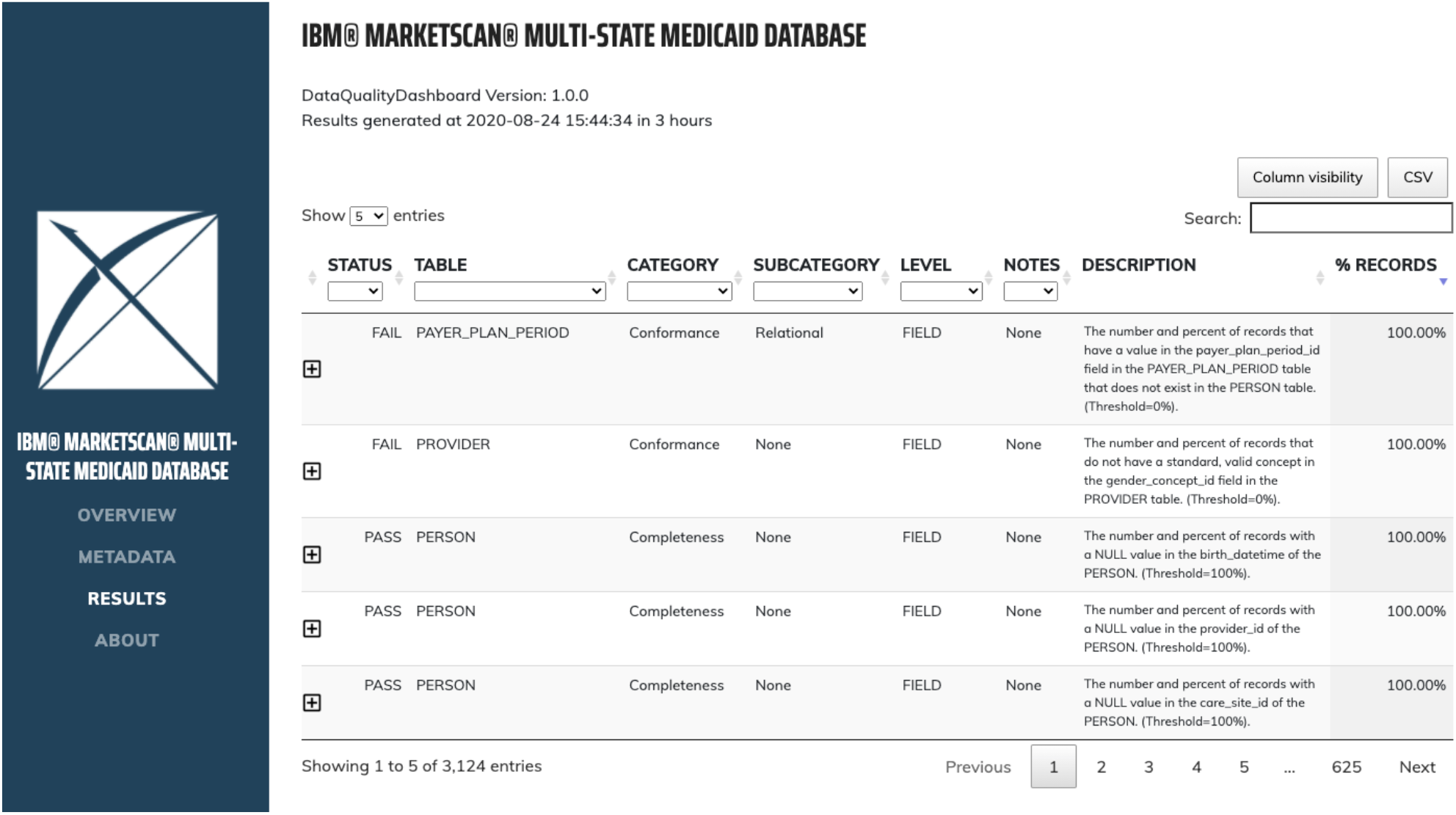
DQD results page with all data quality checks run and their outcomes.

One of the major findings from this exercise showed a high SCRC (19.20%) and high SVC (23.34%) for the PROCEDURE_CONCEPT_ID field of the PROCEDURE_OCCURRENCE table. Further investigation found the problem was due to a previously unaccounted for source vocabulary. Unlike most commercial health insurance plans, state Medicaid coverage usually includes routine dental care. These procedures are coded using Current Dental Terminology (CDT). This had not been documented in the ETL so all dental records were being mapped to a PROCEDURE_CONCEPT_ID of zero. To fix this issue the CDT vocabulary was added to the ETL and the ETL was re-run.

The DQD is available on GitHub[31] as a fully executable R[32] package, supporting OMOP CDM versions 5.2.2 and 5.3.1. It works with multiple database management systems including PostgreSQL, Microsoft SQL Server, and Redshift. All documentation can be found at https://ohdsi.github.io/DataQualityDashboard/.

## Discussion

As use of observational health data continues to increase, two of the largest regulatory agencies in the world have established plans for how these data can be used to support data-driven, regulatory decision-making. The U.S. Food and Drug Administration (FDA) published the Framework for FDA’s Real-World Evidence Program in 2018 which details how and in what capacity real world evidence generated from real world data might be evaluated for its potential use to support approvals of new drug indications or to satisfy post-marketing safety studies.[1] Similarly, the Heads of Medicines Agency (HMA) and the European Medicines Agency (EMA) in 2017 initiated a joint Big Data Task Force. A summary report from 2019 describes a strategy to understand observational data such that they might be ready to make use of it in a regulatory capacity.[2] These reports are robust, especially when discussing how real-world evidence supporting drug safety and effectiveness research should be conducted. Both groups agree that data quality is important and should be considered when determining whether a set of data is suitable to answer specific questions. This especially timely given the COVID-19 pandemic and rush to publish any information that may further understanding of the natural history and clinical treatment of the disease.[33]

The Data Quality Dashboard is well-poised to answer the call for higher quality data. It is at the center of the standard quality control framework (figure 1), providing critical insights to data owners that then feed back into the ETL process, ultimately resulting in research-ready data. There is evidence of broad adoption of the DQD at the network level as the European Health Data and Evidence Network (EHDEN)[34] and the National COVID Cohort Collaborative (N3C)[35] both leverage the framework to ensure that participating databases pass critical quality control measures. As these networks conduct research and learn from their data, any quality issues identified during analysis will be incorporated back into the tool. Continuous iteration on the set of data quality checks in coordination with research creates a living system that improves as we advance our understanding. The DQD will also continue to add features and expand the check types it covers. Initial roadmaps include enabling it to be run on a cohort rather than the entire database, extending the check types to include the OMOP vocabulary, incorporating prevalence measures, and evaluating temporal stability.

Other CRNs have built tools and processes for evaluating and managing the quality of data at the network level. The PEDSNet DQ Workflow [36] utilizes a similar software architecture in that it relies heavily on R, is designed to be applied to databases on the OMOP CDM standard (modified slightly for pediatric use) and can be run on multiple different database management systems (PostgreSQL, Oracle, MySQL, SQLite, or SQL Server). PEDSnet is supported by the Patient Centered Outcomes Research Institute (PCORI) and brings together data from various hospitals into one large, pediatric database. This structure is reflected in the DQ Workflow as the coordinating center has more control over how data quality issues are reported and tracked across the network. Sites run the quality assessment, the results of which are then sent to the coordinating center. Results are reviewed by a data scientist who discusses any issues and corrections needed with the site. The Sentinel Initiative [14,15] has a similar system where Data Partners run a data quality study package, results are sent to the Sentinel Operations Center, and the Operations Center reviews and recommends changes to the ETL. [37] Sentinel data quality checks are divided into different levels and each Data Partner must pass all level 1 checks before moving on to level 2, etc. PCORNet has a five-step data curation process that also involves review of the data quality results by a Coordinating Center. This review is done in cycles whereby network partners are expected to correct any model conformance issues before running completeness checks, for example.

OHDSI, in contrast, is a distributed data network with no central coordination of data. Each member is responsible for their own data and ETL processes. This is a decided choice that allows for faster, broader reaching collaboration. The OMOP CDM could of course be used in a distributed network but sharing protocols and analytic code rather than data eliminates the need to take data beyond the firewall of the data holder. Such a system allows far more sites around the world to contribute as there is no longer an issue of data governance. Therefore, to ensure the quality of the OHDSI network, the DQD had to both assess and communicate the quality of a database while taking differences at the source into account. This led to a tool built using a dynamic framework that expands 20 check types into over 3,300 individual checks. Data owners have full control over which checks are run and how they are assessed for failure while the resulting report details every single check, pass or fail, in an easy-to-share way. It is our vision that this becomes the way data quality is shared among networks, with reviewers, and with regulators. The most recent papers published using the PEDSnet, PCORNet, and Sentinel networks [38–40] either do not mention how the data used were assessed for quality or they touch on it very briefly. Instead, with a tool like the DQD, a JSON file(s) can be shared as additional material to a publication detailing all data quality checks that were run on the database in which the study was executed. This level of transparency is unprecedented in current literature but is necessary in an era where we are asked to trust evidence generated by the scientific community with little to no insight into the data used.

The Data Quality Dashboard has established a new way to garner trust in real-world data. Transparently communicating how well CDM standardized databases adhere to a set of quality measures adds a crucial piece that is currently missing from observational research. Assessing and improving the quality of our data will inherently improve the quality of the evidence we generate.

## Supporting information

results_cdm_mdcd

## Data Availability

The Data Quality Dashboard R package is available on github as is a publicly available instance of the data quality results for IBM MarketScan® Multi-State Medicaid.

https://github.com/OHDSI/DataQualityDashboard/

https://data.ohdsi.org/DataQualityDashboardMDCD/

## APPENDIX

**Table 1:** Data quality check types (ideas) by context and category, also available here: https://ohdsi.github.io/DataQualityDashboard/articles/CheckTypeDescriptions.html

| Check Name | Check Description | Kahn Category |
| --- | --- | --- |
| measurePerson<br>Completeness | *The number and percent of persons in the CDM that do not have at least one record in the @cdmTableName table* | Completeness |
| cdmField | *A yes or no value indicating if all fields are present in the @cdmTableName table as expected based on the specification.* | Conformance |
| isRequired | *The number and percent of records with a NULL value in the @cdmFieldName of the @cdmTableName that is considered not nullable.* | Conformance |
| cdmDatatype | *A yes or no value indicating if the @cdmFieldName in the @cdmTableName is the expected data type based on the specification.* | Conformance |
| isPrimaryKey | *The number and percent of records that have a duplicate value in the @cdmFieldName field of the @cdmTableName.* | Conformance |
| isForeignKey | *The number and percent of records that have a value in the @cdmFieldName field in the @cdmTableName table that does not exist in the @fkTableName table.* | Conformance |
| fkDomain | *The number and percent of records that have a value in the @cdmFieldName field in the @cdmTableName table that do not conform to the @fkDomain domain.* | Conformance |
| fkClass | *The number and percent of records that have a value in the @cdmFieldName field in the @cdmTableName table that do not conform to the @fkClass class.* | Conformance |
| isStandardValid<br>Concept | *The number and percent of records that do not have a standard, valid concept in the @cdmFieldName field in the @cdmTableName table.* | Conformance |
| measureValue<br>Completeness | *The number and percent of records with a NULL value in the @cdmFieldName of the @cdmTableName.* | Completeness |
| standardConcept<br>RecordCompleteness | *The number and percent of records with a value of 0 in the standard concept field @cdmFieldName in the @cdmTableName table.* | Completeness |
| sourceConcept<br>RecordCompleteness | *The number and percent of records with a value of 0 in the source concept field @cdmFieldName in the @cdmTableName table.* | Completeness |
| sourceValue<br>Completeness | *The number and percent of distinct source values in the @cdmFieldName field of the @cdmTableName table mapped to 0.* | Completeness |
| plausibleValueLow | *The number and percent of records with a value in the @cdmFieldName field of the @cdmTableName table less than @plausibleValueLow.* | Plausibility |
| plausibleValueHigh | *The number and percent of records with a value in the @cdmFieldName field of the @cdmTableName table greater than @plausibleValueHigh.* | Plausibility |
| Plausible<br>TemporalAfter | *The number and percent of records with a value in the @cdmFieldName field of the @cdmTableName that occurs prior to the date in the @plausibleTemporalAfterFieldName field of the @plausibleTemporalAfterTableName table.* | Plausibility |
| Plausible<br>DuringLife | *If yes, the number and percent of records with a date value in the @cdmFieldName field of the @cdmTableName table that occurs after death.* | Plausibility |
| plausibleValueLow | *For the combination of CONCEPT_ID @conceptId (@conceptName) and UNIT_CONCEPT_ID @unitConceptId (@unitConceptName), the number and percent of records that have a value less than @plausibleValueLow.* | Plausibility |
| plausibleValueHigh | *For the combination of CONCEPT_ID @conceptId (@conceptName) and UNIT_CONCEPT_ID @unitConceptId (@unitConceptName), the number and percent of records that have a value higher than @plausibleValueHigh.* | Plausibility |
| plausibleGender | *For a CONCEPT_ID @conceptId (@conceptName), the number and percent of records associated with patients with an implausible gender (correct gender = @plausibleGender).* | Plausibility |

## SUPPLEMENTARY MATERIAL

The file results_cdm_mdcd.json has the full data quality results for IBM Marketscan® Multi-State Medicaid.

## Footnotes

1 https://github.com/ohdsi/achilles

